# TRUSTING: An International Multicenter Observational Study of Speech-Based Relapse Prediction in Psychosis Using Explainable AI

**DOI:** 10.1101/2025.11.14.25339774

**Authors:** Roya M. Hüppi, Lucía Bautista, Giacomo Cecere, Sandra A. Just, Sanne Koops, Musarrat Hussain, Enrico Tedeschi, Stephan Benke-Bruderer, Emre Bora, John Lyne, Stefan Kaiser, Elodie Sprüngli-Toffel, Matthias Kirschner, Karl Øyvind Mikalsen, Lars Ailo Bongo, Erik Van der Eycken, Filip Španiel, Brita Elvevåg, Iris E. Sommer, Wolfram Hinzen, Philipp Homan

## Abstract

**Introduction:** The course of psychotic disorders typically involves relapses. Early warning signs vary between individuals and are difficult to detect in clinical practice, especially in outpatient settings. Speech provides a quantitative clinical marker for detecting such early warning signs. The EU Horizon project TRUSTING (A TRUSTworthy speech-based AI monitoring system for the prediction of relapse in individuals with schizophrenia) aims to develop and evaluate a speech-based monitoring system for predicting imminent psychotic relapses. The study will examine the potential for prospective relapse prediction, and feasibility and usability of the monitoring system.

**Methods and analysis:** In this multicenter observational study, *n* = 240 remitted and at-risk-of-relapse adults with psychotic disorders and a comparison group with *n* = 120 healthy participants (matched by age and sex) will be examined at six sites and in six different languages (German, French, Dutch, English, Czech, and Turkish). The follow-up period is 6 months. The TRUSTING smartphone app will be used to collect weekly voice recordings through speech tasks; information on medication adherence, substance use, mood, anxiety, and sleep quality; and motor data from a tapping task. Primary endpoints encompass model performance for relapse prediction, user adherence, transcription quality, usability of recordings, and overall system usability. The primary analysis of user adherence, transcription quality, usability of recordings, and overall system usability will be an unadjusted description of the respective proportions using 95% Wilson confidence intervals. Regarding relapse prediction, the predictive value of the risk estimates for relapse occurrence will be assessed using the area under the receiver operating characteristic curve. Exploratory analysis will be performed on potential speech-based markers associated with relapse risk.

**Ethics and dissemination:** This study has been approved by swissethics (BASEC number: 2025-01177). Findings from this project will be disseminated through peer-reviewed journal publications and presentations at relevant scientific conferences, as well as public events related to mental health.

**ARTICLE SUMMARY:** *Strengths and limitations of this study:*

– International multicenter study spanning six sites, six languages, and five countries, enabling evaluation of the cross-linguistic generalizability of speech-based relapse prediction models in psychosis.
– Human oversight enabling head-to-head comparison between human judgment and machine-generated predictions of relapse risk and ensuring the study’s safety and trustworthiness.
– Involvement of people with lived experience of psychosis in both study and system design.
– Inclusion of a matched control group to study intra- and interindividual variations in speech features over multiple measurements.
– Insights into the feasibility of implementing artificial intelligence (AI)-based transcription and speech analysis in routine mental healthcare, and exploration of novel speech-based markers associated with relapse risk to enhance prediction, understanding, and prevention of relapse in the future.

## INTRODUCTION

Psychosis is a functionally disruptive symptom that occurs across a wide range of conditions.[1] It is characterized by hallucinations, delusions, disorganized behavior, and formal thought disorder,[2] and typically shows a waxing and waning course with remissions and relapses which can be deeply distressing for those affected.[3, 4] Psychosis is a defining feature of schizophrenia spectrum disorders but also commonly presents in bipolar disorder.[1] In the recent World Mental Health Report[4] from the World Health Organization (WHO), the acute phase of schizophrenia was considered the most impairing state across all health conditions. It is estimated that individuals in this state experience only one-fifth of the health and functional capacity of someone in full health. But even in its residual phase, schizophrenia remains the tenth most impairing health state.[4]

One effective method to prevent relapse and concomitant rehospitalization in psychosis is maintenance treatment with antipsychotic medication.[5] However, antipsychotic therapy is associated with serious but variable[6] side effects including cardiovascular disease,[7] sudden cardiac death,[8, 9] weight gain,[10, 11] diabetes,[12] emotional blunting,[13] and sexual dysfunction.[14] Due to these adverse effects, maintenance treatment is increasingly viewed as unacceptable by some individuals with psychosis,[15] leading to high rates of medication discontinuation, with associated increases in relapse risk.[16, 17] One popular alternative for maintenance treatment is targeted or intermittent treatment, in which antipsychotics are administered only when relapse is considered imminent.[18] This highlights the need for relapse prevention strategies that do not rely on continuous antipsychotic treatment[19] and for methods that can accurately identify pre-relapse periods, enabling timely intervention to prevent further deterioration.[20]

Previous research has shown that implementing individualized relapse prevention programs including symptom monitoring, psychoeducational, psychosocial, and/or psychological interventions could also effectively prevent relapse and rehospitalization.[21, 22] Further, the number of rehospitalization days could be reduced with a technology-enhanced relapse prevention program including a web-based prescriber decision support system.[23] In this context, early warning signs, such as changes in sleep patterns or mood, have been used to identify impending relapse,[21] since they are detectable some five weeks before hospitalization.[24] However, when the assessment of early warning signs is based on reports from individuals with psychosis, they show only modest sensitivity and specificity in relapse prediction.[25, 26] Therefore, a marker is needed that is conceptually and temporally related to relapses, sensitive to predict their occurrence, and objectively measurable.[20]

Language has emerged as a promising candidate for such a marker.[20] In individuals with psychotic disorders, language can be altered in various ways, including its content, amount, structure, and acoustic patterns,[2, 27, 28] and is associated with neurobiological abnormalities [29], such as disruptions in language-related white matter tracts. [30–32] In clinical practice, language and speech serve as a fundamental source of data for diagnosis and treatment, and language aberrations might even influence therapeutic alliance and treatment efforts towards recovery.[33] Notably, conceptual disorganization as expressed through speech can be an early sign of relapse,[34] becoming especially prominent in the 2–4 weeks preceding the relapse.[35] Yet, despite careful evaluation of both the content and the manner of speech of an individual with psychosis, even experienced clinicians may miss subtle features and can be influenced by subjective judgments.[36] In addition, most clinical settings allow monthly clinical appointments at best for individuals with psychosis in remission, restricting the opportunity for timely relapse prediction. This challenge creates an opportunity for algorithms based on natural language processing (NLP) or spoken language processing (SLP) to provide quantifiable, clinically relevant insights with objective measurements by offering consistency and eliminating human bias.[37, 38] While NLP-/SLP-derived features have been used to classify individuals with psychosis versus healthy individuals,[39–42] predict psychosis symptom severity,[43] track psychosis symptoms over time,[44] and predict conversion from a high-risk state to psychosis with good to excellent accuracy,[39, 45, 46] they have not yet been used to investigate the prediction of psychosis relapse. Further, there is a need to examine NLP/SLP markers across different populations[47] and languages, since the semantic and acoustic profile of schizophrenia has shown reduced cross-linguistic generalizability in previous research.[28, 48, 49]

Incorporating the collection of speech data into app-based symptom monitoring could be another opportunity.[50] Two recent studies focused on the feasibility of using smartphone applications for relapse prevention and symptom monitoring, and demonstrated that users rated data collection over a 12-month period as feasible. However, user adherence markedly differed between the studies.[51, 52] The usability and feasibility of a monitoring system collecting speech data for relapse risk estimation in individuals with psychosis could provide an adequate non-intrusive way to monitor these individuals,[20], but has not been investigated so far.

Against this background, the present research project aims to investigate the potential of automated, artificial intelligence (AI)-based transcription and speech analysis within a digital monitoring system to estimate relapse risk in individuals with psychosis. We hypothesize that the monitoring system will achieve an area under the receiver operating characteristic curve (AUC) of at least 0.70[34] for prognostic prediction in individuals with psychosis, assuming a 6-month relapse prevalence of 26% (weekly probability of 3%).[53] In line with the goals of explainable AI, the aim is for the underlying models to be interpretable and theoretically motivated, satisfying recommendations for clarity on how, why and what we are measuring.[48] To enhance the trustworthiness of the monitoring system, human involvement will be crucial to this study. Humans-in-the-loop (HITL) will ensure that the data quality is high, and the machine-generated predictions of relapse risk will be compared head-to-head with human judgement. Furthermore, the needs of people with lived experience of psychosis have been and will continue to be actively incorporated into the study by involving them in designing both the study and the monitoring system.

Aiming for a longitudinal, multilingual database, we will collect speech, self-report, and motor data on a weekly basis across six languages (German, French, Dutch, English, Czech, and Turkish) using the TRUSTING smartphone application. Consecutively, we will aim to assess the feasibility and usability of multilingual, smartphone-based data collection in a naturalistic setting. We expect that 70% of users will be adherent, and therefore complete at least 33% of their tasks.[54, 55] High-quality transcripts, i.e., transcripts with a word error rate (WER) of at most 35%, are expected in 60% of participants.[56] The proportion of participants producing usable audio recordings—recordings in which the question is interpreted correctly, and the audio is of sufficient quality—is conservatively expected to be 70%.[57] In addition, we expect 75% of participants to rate the TRUSTING app as usable, thus rating it with a score of at least 70[53] on the System Usability Scale (SUS)[54]. Finally, in line with the exploratory nature of the study, we aim to explore novel relapse markers that might ameliorate the current understanding of relapse mechanisms and may be useful as predictive or diagnostic tools in future clinical practice.

## METHODS AND ANALYSIS

This protocol adheres to the Strengthening the Reporting of Observational Studies in Epidemiology (STROBE) guidelines.[58]

### Study Summary

Table 1 provides a summary of the “A TRUSTworthy speech-based AI monitoring system for the prediction of relapse in individuals with schizophrenia (TRUSTING)” study information.

**Table 1:** Study summary.

|  |  |
| --- | --- |
| Study title | TRUSTING: An International Multicenter Observational Study of Speech-Based Relapse Prediction in Psychosis Using Explainable AI |
| Short title | TRUSTING Observational Study |
| Study design | Observational, prospective, longitudinal, exploratory, international, multicenter |
| Study participants | Individuals at risk of psychosis relapse recruited from clinical settings during a stable phase of partial or full remission and healthy individuals |
| Planned sample size | 240 individuals with psychosis and 120 healthy individuals |
| Study sites | 6 sites covering 6 languages (German, French, Dutch, English, Czech, Turkish) in 5 countries (Switzerland, the Netherlands, Ireland, the Czech Republic, Turkey) |
| Start date | *tbd* |
| Planned study period | 12 months |
| Follow-up duration | 6 months |
| Ethics / registration number | This study has been approved by swissethics (BASEC number: 2025-01177). |
| Aims | To investigate the potential of automated, AI-based transcription and speech analysis within a digital monitoring system to estimate relapse risk in individuals with psychosis.<br>To collect speech, self-report, and motor data across six languages (German, French, Dutch, English, Czech, and Turkish) using the TRUSTING app, and to evaluate the feasibility and usability of data collection.<br>To explore novel relapse markers that might enhance the current understanding of relapse mechanisms and may be useful as predictive or diagnostic tools in future clinical practice. |
| Project website | https://trusting-project.eu/ |

### Study Setting and Design

This international multicenter research project will be conducted across six sites, with data collected in six different languages. In Switzerland, both German data (University of Zurich) and French data (University of Geneva) will be gathered. Dutch data will be collected in the Netherlands (University Medical Center Groningen), English data in Ireland (RCSI University of Medicine and Health Sciences), Czech data in the Czech Republic (National Institute of Mental Health), and Turkish data in Turkey (Dokuz Eylul University). The study follows an observational, prospective, longitudinal, exploratory design. The overall planned project duration is 12 months, commencing once ethical approval and all required agreements are secured. The recruitment and screening phase is expected to last approximately 6 months, with a follow-up duration of 6 months. Ethical approval has been sought for an extended overall study and follow-up period, allowing the study duration to be extended if additional time is needed to recruit a sufficient number of participants and collect adequate data.

After providing the participants with comprehensive information on the study, written informed consent of the participants will be obtained before enrolling them in the study. Data collection will start with an on-site clinical assessment (T0), followed by a 6-month period of weekly data acquisition using the TRUSTING app. Participants will provide voice recordings through speech tasks; self-reported information on medication adherence, substance use, mood, anxiety, and sleep quality; and motor data from a tapping task. The TRUSTING app is an adaptation of the *delta* Mental Status Exam application.[59] Participants will be instructed to complete the weekly sessions in a private setting. A follow-up clinical assessment will be conducted on site after 6 months (T1). The study timeline is visualized in Figure 1. Figure 3 provides details on the variables assessed at each step.

**Figure 1:**
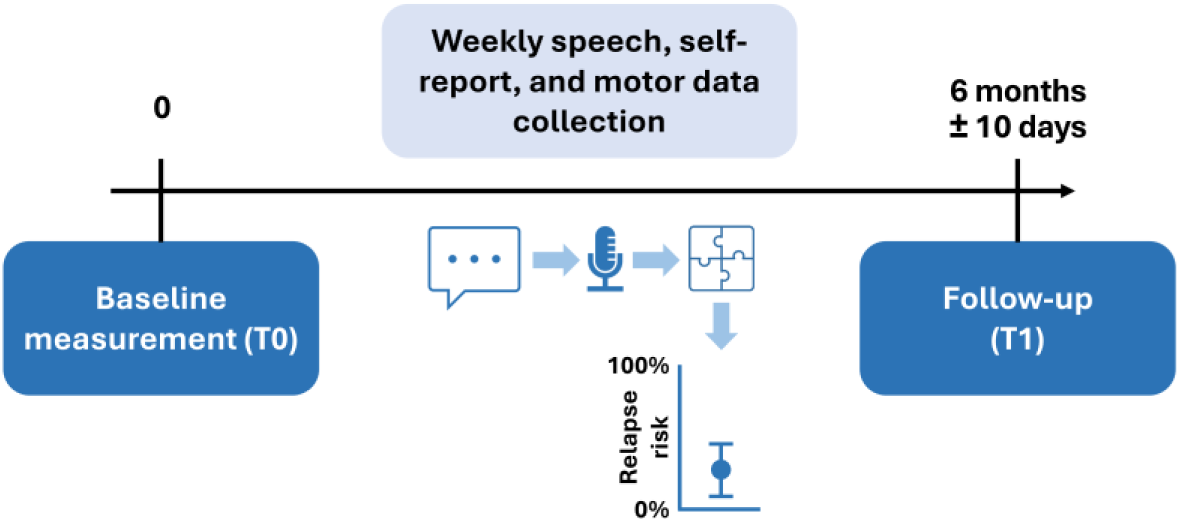
Study timeline including the on-site clinical assessments (T0 and T1) and weekly speech, self-report, and motor data recordings from which relapse risk will be estimated.

#### Data Preprocessing

After a participant has completed a session in the TRUSTING app, their data will be transferred to be stored securely within a research platform for sensitive data (TSD). The TRUSTING research partner in Norway at UiT – The Arctic University of Norway has created the technical infrastructure for the project, including the backend data management and processing system within TSD, which is hosted at the University of Oslo. Within the TSD environment, speech recordings will be automatically transcribed using AI-based methods and raw scores will be extracted from the tasks.

#### Human Oversight

To ensure data quality and model validity, human oversight will be a key component of this study. HITL will be researchers or clinicians involved in the study at the research site but not involved in the treatment of the participants whose data they evaluate. They will be trained for the task at hand. After the data has been preprocessed within the TSD, a first human-in-the-loop (HITL1) will review the data quality by checking the audio quality and accuracy of the automated transcripts of the audio recordings, correcting transcription errors to ensure accurate subsequent analysis, and flagging any potential problems with the data, such as poor audio quality or other data anomalies. They can but will not be obliged to know the identity of participants.

After the data quality check is completed, a second human-in-the-loop (HITL2) will be involved. The HITL2 must be different to the clinician in charge and will not be allowed to know the identity of the participants they are overseeing. Importantly, they will not have access to the relapse risk estimate provided by the digital monitoring system (see <u>Digital Monitoring System</u>) or any demographic information about or group membership of the participants within the TSD. As a first task, the HITL2 will be presented with plots showing the development of raw response data (medication adherence, recreational drug use, mood, anxiety, sleep quality) across all completed sessions up to the current one. For each raw score, the HITL2 will rate their level of agreement with the statement that the participant’s score of the current session implies a risk (on a five-point Likert scale from disagree to agree). Moreover, the HITL 2 will listen to the speech recordings from the current session and answer two further questions about whether the current speech content or acoustic features indicate risk of relapse (also on a five-point Likert scale of agreement). Next, the HITL2 will provide their own relapse risk estimate for this session (from 0 = no risk to 1 = high risk) as well as the confidence level of their risk estimate (from 0 = no confidence to 1 = high confidence). Third, for each session the HITL2 will estimate whether the participant belongs to the group of individuals with psychosis or the healthy control group. Finally, they will suggest a clinical decision based on their own relapse risk assessment. This recommendation will be recorded for later analysis but will not be shared with the clinician in charge, as this study is purely observational. Apart from the data presented in the TSD annotation interface for the current session, the HITL2 also has the option to access evaluations from past sessions.

#### Digital Monitoring System

In addition to the human risk assessment, the TRUSTING project involves the development of a digital monitoring system that provides a relapse risk estimate for each person at every session. The estimate will be based on the speech, self-report and motor data that was collected from individuals with psychosis and healthy individuals via the TRUSTING app, automatically preprocessed, and checked by the HITL1. Alongside the absolute task values, the predictive model will include information on within-individual changes of variables over time to reduce confounding from stable interindividual differences and to thereby improve the performance of the predictive model.[56]

For each task, a single risk score will be calculated, ranging from 0 (no risk) to 1 (high risk). If data from an individual task is missing, that task will be assigned the highest possible risk score of 1, while completed tasks will retain their calculated values. As the strength of evidence for the association with relapse risk prediction varies between tasks, different weights will be assigned to each task based on expert knowledge and the literature on relapse risk in psychosis. Initially, equal weights will be assigned to each task, with the exception that tasks related to antipsychotic medication adherence and the use of recreational drugs will be given double weight due to their clinical relevance.[60] These initial weights will serve as a starting point and will be refined or validated through analysis using synthetic data as well as data from the observational study.

The final risk score for each session will be calculated by aggregating the task risk scores and their respective weights. If the data for an entire session is missing, the corresponding week will be assigned the highest possible risk score of 1. Risk scores will be mapped to one of three risk categories, as shown in Table 2. Accumulative score ranges of 0–0.2 indicate low risk, > 0.2–0.6 indicate medium risk, and > 0.6 indicate high risk. These ranges were selected by experts to balance the need to avoid both false positives and false negatives. As part of striving for explainable AI, explanation generation through visualizations is integrated into the system to make outputs interpretable for researchers.

**Table 2:** At the end of each session, risk scores ranging from 0 to 1 will be computed for each participant by aggregating their weighted task risk scores for that complete session. The final risk score for each session corresponds to three relapse risk categories: low, medium, and high risk. This is the final output of the digital monitoring system per person per session.

| Final score | Final prediction |
| --- | --- |
| 0–0.2 | Low risk |
| > 0.2–0.6 | Medium risk |
| > 0.6 | High risk |

#### Exploratory Research

In addition to the relapse risk assessment by the HITL2 and the monitoring system, an exploratory research branch will be incorporated in the study to identify new speech markers associated with relapse. This will involve the search for additional speech features beyond previously established markers to improve understanding of relapse mechanisms. Newly discovered relapse markers could potentially serve as predictive or diagnostic tools for future clinical use. See Figure 2 for a broad overview of the study.

**Figure 2:**
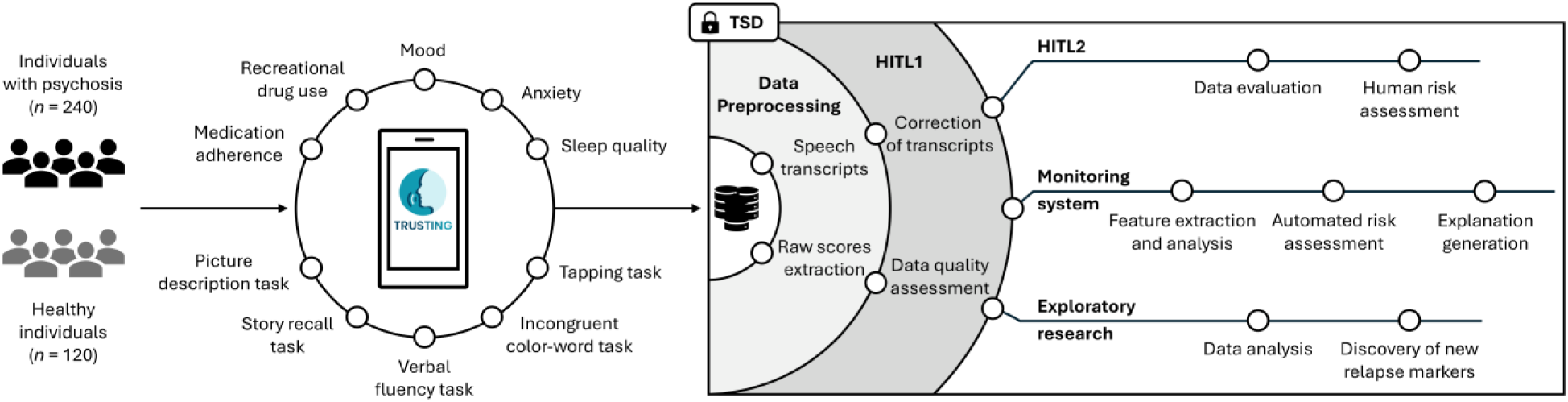
Broad overview of the study. The TRUSTING smartphone app will be used to collect weekly voice recordings through speech tasks (picture description, story recall, verbal fluency, and incongruent color-word task), self-reported information on medication adherence, recreational drug use, mood, anxiety, and sleep quality as well as motor data from a tapping task from individuals with psychosis and healthy individuals. All data will be transferred into a secure data storage (TSD), where they will be automatically preprocessed. Humans-in-the-loop 1 (HITL1) will correct transcripts and assess data quality. Humans-in-the-loop 2 (HITL2) will evaluate the data and assess relapse risk. The monitoring system will extract and analyze relevant features from the data and provide an automated risk assessment. Explanation generation is integrated into the system to make outputs interpretable for researchers. An exploratory research branch will focus on identifying new markers associated with relapse by analyzing features beyond those already established.

### Recruitment

Individuals at risk of psychosis relapse will be recruited from clinical settings during a stable phase of partial or full remission. Depending on the site, eligible participants may be inpatients approaching discharge or outpatients receiving ongoing care. Healthy individuals will be recruited via community outreach, advertisements, and volunteer registries associated with universities and hospitals. Individuals expressing interest in the study will undergo an initial prescreening to ensure they fulfill the <u>Inclusion Criteria</u> and do not meet any of the <u>Exclusion Criteria</u>.

#### Inclusion Criteria

To be included in the study, all participants have to be: (1) between 18 and 65 years old; (2) able to provide informed consent; (3) able to comply with the study schedule and procedures; (4) native or equivalently fluent in speaking and understanding either German, French, Dutch, English, Czech, or Turkish, depending on the site; (5) able to have access to a smartphone; (6) capable of using a mobile app for speech-based data collection; and (7) able and willing to understand the purpose and the details of this study. For individuals with psychosis, inclusion criteria additionally will be: (1) a diagnosis of a psychotic disorder (schizophrenia, schizoaffective disorder, schizophreniform disorder, acute psychosis, bipolar disorder with psychotic symptoms, or psychosis not otherwise specified); (2) first visit during remission phase (baseline measurement); and (3) current positive symptoms rated 3 (mild) or less on all of the following Brief Psychiatric Symptom Scale (BPRS) [61] items: hallucinatory behavior, unusual thought content, conceptual disorganization.

#### Exclusion Criteria

For individuals with psychosis, (1) severe comorbid speech disorders (aphasia or severe stuttering) that prevent adequate speech recording will lead to exclusion. Psychiatric or somatic comorbidity and harmful use of alcohol or other substances will not result in the exclusion of individuals with psychosis, so that the sample will reflect the general population of individuals with psychosis and the study endpoints will be generalizable. Healthy participants’ exclusion criteria will be: (1) any major psychiatric or medical condition; and (2) harmful use of alcohol or other substances.

#### Matching Strategy

Aiming for demographic comparability between groups, healthy participants will be selected to match individuals with psychosis based on key characteristics such as age and sex in each site. The matching will be performed either exactly or within defined age ranges (i.e., 18‒40 years and 41‒65 years). Initially, recruitment will adopt a broader approach to establish a pool of eligible healthy volunteers. As enrollment of individuals with psychosis advances, recruitment of healthy participants will become more targeted, prioritizing those who closely match the characteristics of newly enrolled individuals with psychosis. By refining the matching process over time, we aim to optimize comparability between the two groups while maintaining recruitment efficiency. Additionally, the recruitment strategy will include a minimum representation of 40% female participants (maximum 60%) and 40% of participants aged over 40 years (maximum 60%) to allow for meaningful analysis and enhance the generalizability of findings.

#### Participant Retention and Follow-up Strategies

To minimize withdrawal, clear communication during informed consent will ensure participants understand the study’s purpose, benefits, and data confidentiality. Regular check-ins and open communication channels will help address concerns and reinforce engagement. Flexible participation options, such as remote follow-up and temporary pauses, will reduce burden, while pseudonymization will help alleviate privacy concerns. Reminder systems and appropriate incentives will further support retention. To reduce loss to follow-up, proactive participant retention strategies will be implemented, including regular follow-up via phone or email, documentation of contact attempts, and tracking reasons for participant attrition. All participants, including those who withdraw or are lost to follow-up, will be fully documented to maintain transparency in the study’s analysis. The detailed participant timeline is presented in Figure 3.

**Figure 3:**
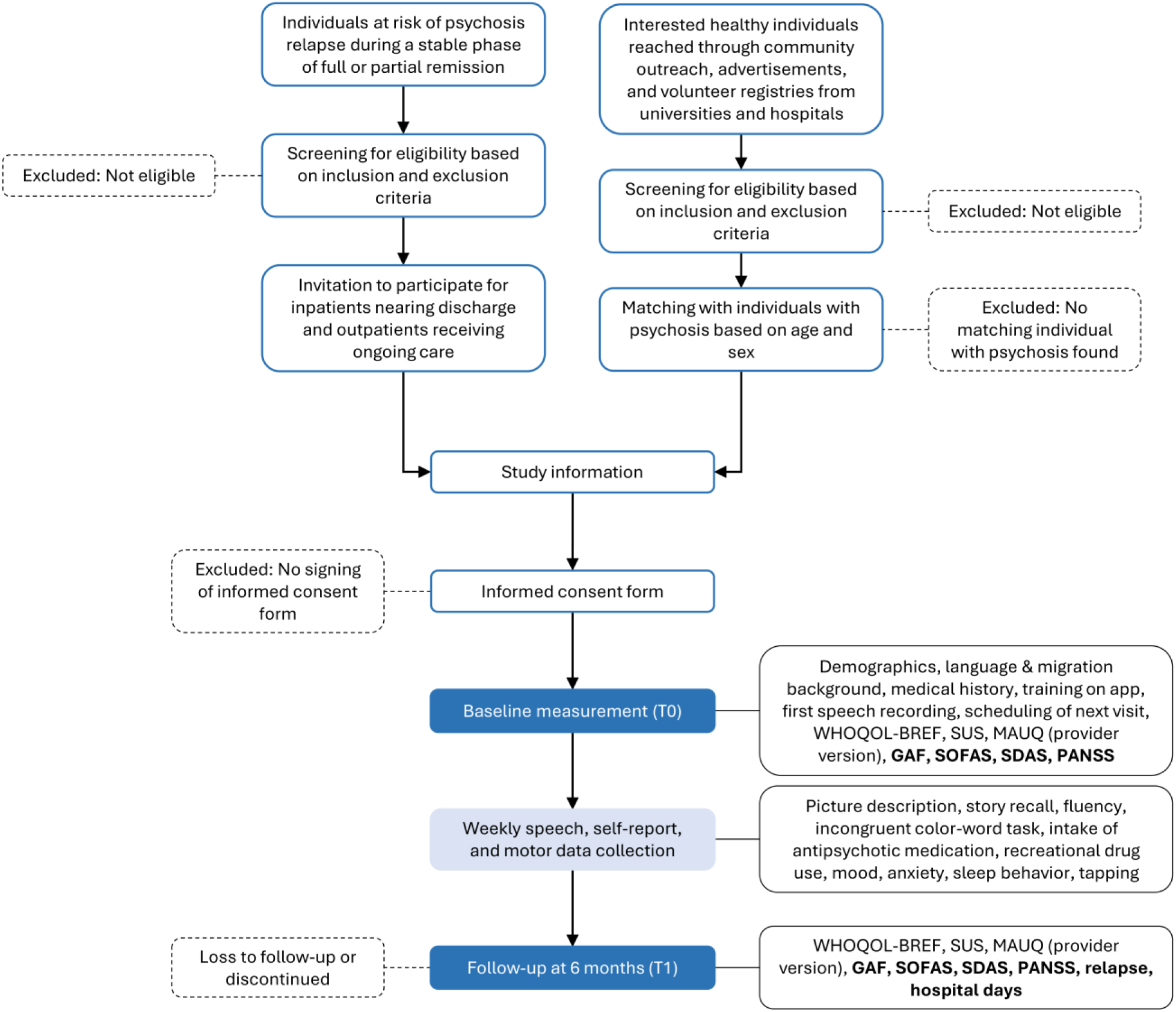
Detailed participant timeline covering the recruitment process, provision of study information, obtaining informed consent, on-site assessments and weekly speech, self-report, and motor data collection. Variables written in bold are only assessed in individuals with psychosis. WHOQOL-BREF = Short version of the World Health Organization Quality of Life Assessment; SUS = System Usability Scale; MAUQ = mHealth App Usability Questionnaire; GAF = Global Assessment of Functioning; SOFAS = Social and Occupational Functioning Assessment Scale; SDAS = Social Dysfunction & Aggression Scale; PANSS = Positive and Negative Syndrome Scale.

### Variables

#### Baseline Data and Confounders

Data pertaining to sociodemographic and clinical variables will be collected to characterize the study population and to inform statistical adjustment in subsequent analyses. These variables include age, sex, education, language, migration background, medical history, diagnoses, concomitant medication, medication adherence, substance use, mood, anxiety, motor data from a tapping task, and sleep quality. A subset of these—specifically sex, age, education, language, medication adherence, and substance use—will be considered as potential confounders in predictive models. This data will be obtained during the on-site assessments and through weekly app recordings.

#### Primary Outcomes

The primary outcomes we will assess are the performance of the digital monitoring system’s relapse risk estimates for relapse prediction, user adherence, transcription quality, usability of recordings, and the overall system usability in individuals with psychosis.

##### Performance of the Digital Monitoring System for Relapse Prediction

The digital monitoring system will provide a weekly relapse risk estimate for each participant, represented as a continuous value between 0 and 1, where higher values indicate a higher predicted risk of relapse. To assess the predictive performance of the monitoring system, this risk estimate will be compared to the actual relapse outcome, based on observed clinical data during the following week. Relapse will be defined as hospitalization, since hospitalization is often a sufficient indicator of relapse,[62] and will be coded as a binary variable (0 = no relapse, 1 = relapse). The performance of the monitoring system for relapse prediction in individuals with psychosis will be assessed by the area under the receiver operating characteristic curve (AUC). The AUC reflects the probability that a randomly selected individual with psychosis who experiences a relapse will have a higher estimated risk score than a randomly selected individual with psychosis who does not relapse. In addition to the AUC, various threshold values of the monitoring system’s risk estimate will be explored to determine optimal cut-off points for classifying individuals with psychosis as high or low risk for relapse.

##### User Adherence

User adherence refers to the proportion of users who adhere to their allocated tasks. Each user’s task completion will be automatically assessed. A user will be considered adherent if they complete at least 33% of their tasks across all sessions.[54, 55]

##### Transcription Quality

Transcription quality is defined as the proportion of users for whom high-quality transcripts are produced. Transcription quality will be measured by the WER between automated speech recognition and human-corrected transcripts. A user with an overall WER of at most 35% will be regarded as having high-quality transcripts.[56]

##### Usability of Recordings

Usability of recordings relates to the proportion of speech recordings that can be used for speech analysis. Usability of each record will be assessed by HITL reviewers based on the following three questions: has the user has interpreted the question correctly; is the audio (at least partially) audible; and is the audio (at least partially) comprehensible. The three questions must be answered with “yes” for the recording to be rated as usable.

##### Overall System Usability

For the overall system usability, the proportion of users rating the TRUSTING app as usable will be assessed. Usability will be evaluated using the System Usability Scale (SUS),[63] providing a standardized usability score (range 0–100) based on a 10-item questionnaire. Each item will be rated using a 5-point Likert scale from 0 (strongly disagree) to 4 (strongly agree). The sum of the score must be multiplied by 2.5 to obtain the overall score. The monitoring system will be deemed usable with a score of at least 70.[64] The SUS is effective and efficient,[65] and demonstrates strong face validity and very high reliability (Cronbach’s α = 0.91).[64]

#### Secondary Outcomes

##### Relapse Rate

The proportion of individuals with psychosis experiencing at least one relapse (defined as rehospitalization) during the study duration will be assessed as the relapse rate.

##### Performance of the HITL for Relapse Prediction

Equal to the evaluation of the performance of the digital monitoring system for relapse prediction, the performance of the HITL relapse risk estimates for relapse prediction in individuals with psychosis will be assessed by the AUC. Thresholds for relapse prediction will again be explored as part of the analysis.

##### Human-Machine Agreement in Relapse Risk Estimation

The level of agreement between the HITL and the monitoring system’s relapse risk estimates will be calculated to enable head-to-head comparison between human judgment and machine-generated predictions.

##### Risk Category-Based Analysis of Relapse Outcomes

To further evaluate their relationship with the incidence of relapse, the HITL and the monitoring system’s relapse risk estimates will be transformed from continuous scores (from 0 to 1) into categorical risk levels (low risk: 0–0.2, medium risk: > 0.2–0.6, high risk: > 0.6). Subsequently, descriptive analysis of the proportion of relapses in those categories will be performed.

##### Quality and Usability Measures in Healthy Participants

User adherence, transcription quality and usability of recordings as well as the overall system usability will be assessed in healthy participants and then compared to the equivalent values in individuals with psychosis.

##### Provider-Perceived Usability

To evaluate the provider-perceived usability and usefulness of the app, the provider version of the mHealth App Usability Questionnaire (MAUQ)[66] will be used. Scores will be reported by domain and overall. The MAUQ demonstrates high internal consistency of the three subscales (Cronbach’s α between 0.83 and 0.90) and the entire questionnaire (Cronbach’s α = 0.93). It further shows strong criterion and construct validity as the MAUQ overall score is correlated with the Post-Study System Usability Questionnaire (PSSUQ, *r* = 0.84) and the SUS (*r* = 0.64), two commonly used usability questionnaires.[66] MAUQ scores will be assessed by clinicians in charge at baseline and after 6 months.

##### Speech- and Text-Based Features

Speech-based features will be extracted from weekly speech recordings using tools such as Prosogram or openSMILE. In addition, robust text-based features will be derived from transcripts using common Python NLP libraries like NLTK and spaCy. WordNet will be used to assess the correctness of responses where appropriate (fluency task, story recall task, and incongruent color-word task). The speech- and text-based features will be exploratively compared between individuals with psychosis and healthy participants.

##### Quality of Life

Quality of life will be assessed at baseline and after 6 months in both individuals with psychosis and healthy participants using the short version of the World Health Organization Quality of Life (WHOQOL)[67] assessment: the WHOQOL-BREF[68]. The WHOQOL-BREF demonstrates good internal consistency (Cronbach’s α between 0.66 and 0.84), test-retest reliability for items (*r* = 0.56–0.84) and domains (*r* = 0.66–0.87), discriminant validity, and content validity.[68]

##### Global, Social and Occupational Functioning

The Global Assessment of Functioning (GAF)[69] will be used to measure how much a person’s symptoms affect their day-to-day life on a scale of 0 to 100. An advantage of GAF is its simplicity.[70] The Social and Occupational Functioning Assessment Scale (SOFAS)[71] will provide a global rating of current social and occupational functioning with scores ranging from 0 to 100. It differs from the GAF by assessing social and occupational functioning independently of the individual’s overall psychological symptom severity.[72] GAF and SOFAS scores will be recorded at baseline and after 6 months, but only in individuals with psychosis.

##### Psychopathology and Symptom Severity

To measure the prevalence of positive and negative syndromes in schizophrenia, the Positive and Negative Syndrome Scale (PANSS)[73], one of the most widely used instruments in schizophrenia research, will be used. The PANSS is a 30-item instrument including a positive (7 items), a negative (7 items) and a general psychopathology scale (16 items). Each item will be rated using a 7-point Likert scale from 1 (absent) to 7 (extreme). The PANSS demonstrates high interrater reliability with mean correlation values between 0.83 and 0.87, acceptable to good internal consistency (Cronbach’s α between 0.73 and 0.83), and moderate to strong test-retest reliability (*r* = 0.60–0.80).[73, 74] It will be measured in individuals with psychosis at baseline and after 6 months.

##### Aggression and Risk Behaviors

The rates of self-harm (including suicide, suicide attempts and aggressive incidents) will be assessed with the Social Dysfunction & Aggression Scale (SDAS)[75]. The SDAS demonstrates adequate inter-observer reliability and good internal consistency (Cronbach’s α of 0.79).[75] The rates of self-harm will be collected in individuals with psychosis at baseline and after 6 months.

##### Psychiatric Admissions

The number and duration (in weeks) of psychiatric admissions within the 6-month follow-up period will be recorded for individuals with psychosis.

### Statistical Analyses

Descriptive analyses will include estimates with confidence intervals (CI) as well as graphical representations of the data. The exploratory analysis will mainly rely on generalized linear (mixed) models that will be built in a data-driven manner and their estimates for fixed effects and estimated marginal means. For the fixed effects of the models, Kenward’s method[76] will be used to estimate the degrees of freedom for the calculation of p-values and CIs. When estimated marginal means of factor levels are reported, Wald CIs will be used. Participants’ contributions in the models will be weighted linearly according to their participation duration where appropriate. The following potential confounders will be considered when building the models: sex, age, education, language, medication adherence and substance abuse.

The analysis of the user adherence, transcription quality, usability of recordings, overall system usability and relapse rate endpoints will be an unadjusted description of the respective proportions using 95% Wilson confidence intervals. In the exploratory analysis of these endpoints, we aim to describe as well as find potential differences in the following populations of interests or combinations thereof: participant group (psychosis/healthy), sex, age, and language categories.

Regarding relapse prediction, the predictive value of the risk estimates for relapse occurrence (from the HITL and the monitoring system) will be assessed using receiver operating characteristic (ROC) curves and the corresponding AUC. Potential thresholds for relapse prediction by the HITL and the monitoring system will be explored. To assess the level of agreement between the HITL and the monitoring system’s relapse risk estimates, the Spearman correlation of the actual estimates (ranging from 0 to 1) as well as Cohen’s kappa for categorized predictions (low risk: 0 to 0.2, medium risk: > 0.2 to 0.6, high risk: > 0.6) will be calculated. For a descriptive analysis, the proportion of observed relapses in these categories will be described for the HITL and the monitoring system’s relapse risk estimates. The association of relapse with clinical (symptom severity) and behavioral parameters will be investigated using logistic regression models.

Concerning the recordings, descriptive and graphical analyses of the number of recordings per participant, interpretation of questions and comprehensibility and audibility of audio across individuals with psychosis and healthy participants will be performed. To investigate the behavior of speech-derived features over time, explorative longitudinal models with an emphasis on how these features are influenced by participant group and language will be built. Descriptive analysis will be carried out using graphical representations of the time courses.

Descriptive analyses of the number and duration of psychiatric admissions, the rates of self-harm (including suicide, suicide attempts and aggressive incidents) assessed with the SDAS, as well as GAF, SOFAS, WHOQOL, and PANSS scores will be performed. Models will be built to compare quality of life assessed with the WHOQOL between participant groups and to assess the association with clinical, social, and functional variables.

#### Sample Size

A sample size calculation was performed for each primary outcome. For user adherence, we conservatively expect 70% (desired precision 60% to 80%) of participants to complete ≥ 33% of their tasks,[54] leading to a required sample size of 78 individuals with psychosis. Regarding the transcription quality, we set the WER threshold at ≤ 35% and expect 60% (desired precision 50% to 70%) of participants to meet the threshold.[56] The estimated sample size for transcription quality is 89 individuals with psychosis. Concerning the usability of recordings, we conservatively expect 70% (desired precision 60% to 80%) of speech recordings to be of high quality.[57] This would require 78 individuals with psychosis to reach the desired estimate precision. For the usability of recordings endpoint, each participant can contribute up to 208 recordings. However, we expect a high intra-individual correlation in recording quality, meaning that repeated recordings from the same individual are likely to be similar. To adopt a conservative approach, we assume the worst-case scenario in which repeated recordings provide no additional information beyond the first. Accordingly, we set the required number of participants for this endpoint equal to the number of independent recordings needed—i.e., 78 individuals with psychosis—to ensure adequate precision under this assumption.

Accurately estimating the performance of the monitoring system for relapse prediction may require a very large sample size, since the within-person correlation is unknown. Therefore, we aim to recruit 240 individuals with psychosis across six sites (40 per site); this represents the maximum sustainable number for this observational study. Relapse risk will be predicted weekly using all available data from the individual, even after relapse. Across six months, each individual will contribute up to 26 repeated, correlated weekly predictions, with an optimistic presumed 6-month relapse probability of 26%.[53] We expect an average relapse duration of 3 weeks[77–79] resulting in a weekly relapse prevalence of 3% (optimistically). The anticipated AUC for relapse prediction is conservatively estimated at 0.7, which is comparable to the AUC of 0.72 observed in the cross-site validation of a model predicting which high-risk individuals will develop psychosis.[39] To account for clustering of predictions within participants, we applied a design effect.[66] We further adjusted for an anticipated dropout rate of 33%. Based on a total of 240 individuals with psychosis, the 95% confidence interval of the AUC narrows as within-person correlation decreases. Using the prec_auc() function of the *presize* R package[80], which applies the method of Hanley and McNeil[81], we determined that the maximum tolerable within-person correlation before the lower CI limit of the AUC falls below 0.5 (which would indicate no predictive value) is 0.5.

In addition to the 240 individuals with psychosis, 120 healthy participants (20 per site) will be included in the study. The number of healthy participants is lower than that of individuals with psychosis, as we expect reduced variability and lower uncertainty within this group. Including 20 healthy participants per language is considered sufficient to incorporate language as a variable in the model, following the general rule of thumb for predictor inclusion.[82]

#### Missing Data

The frequency, patterns, and causes (if available) of missing data for the endpoints will be analyzed. The employed statistical models can handle missing data under the missing at random assumption. However, the handling of missing values in the automated risk score computation of the monitoring system is an exception, as described in the section <u>Digital Monitoring System</u>.

## ETHICS AND DISSEMINATION

### Data Management

This research project will be conducted in accordance with the Declaration of Helsinki[83] as well as locally relevant regulations. The electronic data capture (EDC) software secuTrial® will be used for data processing and management. Any study-related speech recordings, self-report data, and motor data collected with the TRUSTING app will be securely stored in the TSD. Both the EDC and TSD provide appropriate safeguards to ensure confidentiality and integrity of the data and follow data protection laws and regulatory standards. Regarding data encryption, pseudonymization will be employed to protect participants’ privacy throughout the research process.

Access to the data will be restricted to authorized personnel only. Summary results will be made available in accordance with applicable ethical and regulatory requirements. Project data will be retained in a pseudonymized format for the required retention period of at least ten years, as by the Swiss Human Research Act (HRA) and Human Research Ordinance (HRO), the European General Data Protection Regulation (GDPR), and the Turkish Personal Data Protection Law (KVKK), as well as applicable ethical approvals. In the event of a participant’s withdrawal from the study, participants can request the deletion of their data directly through the TRUSTING app in accordance with the New Swiss Federal Act on Data Protection (nFADP), GDPR, and KVKK (right to be forgotten).

### Patient and Public Involvement

The Global Alliance of Mental Illness Advocacy Networks-Europe (GAMIAN-Europe), representing the interests and rights of persons affected by mental health conditions (including psychotic conditions), plays a critical role in ensuring the success of the TRUSTING project. GAMIAN-Europe is involved in the study and monitoring system design process, facilitates the dissemination of information and resources related to the project, and ensures effective communication of developments and findings to all stakeholders, particularly individuals with psychosis and the national patient organizations advocating on their behalf.[84] With the assistance of GAMIAN-Europe, we established a user board of representatives with lived experience who co-designed the TRUSTING app together with the UiT research team. This intense collaboration ensures that the app aligns with user needs and perspectives. The board extensively tested the app, provided essential feedback through a survey, and contributed to this study protocol. The public was not involved in the design, conduct, reporting, or dissemination plans of this research.

## ETHICS AND DISSEMINATION

Ethical approval is required for each country involved in data collection, storing, or processing; namely Switzerland, the Netherlands, Ireland, the Czech Republic, Turkey, and Norway. The site in Zurich, Switzerland, will coordinate the observational study as the study sponsor.

In this study, participant data will be analyzed using AI techniques, including NLP, SLP, and predictive modeling of relapse risk. The use of AI will comply with Swiss nFADP and EU GDPR regulations, and, where applicable, the EU Artificial Intelligence Act. All participants will be aware of the use of AI and will be asked to provide explicit informed consent for their data to be used in this manner. Striving for explainable AI, we aim to build fully interpretable and theoretically motivated models, clarifying the object and procedure of measurement, as well as the underlying rationale. AI analysis outputs will be used solely for research purposes to identify trends and patterns, not to inform individual clinical decisions.

This research project has been designed with careful attention to both the scientific value and ethical implications. The results of this project are expected to have significant social and scientific value by advancing understanding of relapse risk in individuals at high risk for psychosis relapse. The findings may contribute to the development of predictive tools for mental health management, benefiting persons, healthcare systems, and the broader community. However, the generalizability of these results will be considered in light of the study’s observational nature and the specific population involved.

The project design and participant procedures have been carefully justified to minimize participant burden by keeping the time and effort demanded of participants to a reasonable level. The overall benefits to science and society, including the potential for early identification of relapse risk, outweigh the minimal burdens placed on participants. Participation in this project is entirely voluntary, forming a fundamental principle of the project. Participants will be informed of their right to withdraw at any time without any consequence. All data will be handled confidentially, with strict adherence to ethical standards for privacy and protection of personal information.

Special attention is given to ethical considerations around incidental findings and surplus information. Should clinically relevant incidental findings arise—information not originally required for the project, but which may impact a participant’s health—appropriate steps will be taken to inform the participant in accordance with established local guidelines, such as the “Guideline for handling incidental findings in medical research” published on swissethics.ch. Participants will be provided with clear and transparent information regarding their right to receive information about incidental findings, including the potential risks and benefits of knowing such findings.

Findings from this project will be disseminated to healthcare professionals and the scientific community through peer-reviewed journal publications and presentations at relevant scientific conferences. The publication of this study protocol is an important first step toward ensuring the visibility and transparency of our research.

A lay summary of the findings will also be published in the Business Administration System for Ethics Committees (BASEC) in Switzerland and will be available in the Swiss national languages relevant to participant recruitment, i.e., German and French. Additionally, the study results will be disseminated through project-related communication channels such as the project website (https://trusting-project.eu), social media platforms, and newsletters. These public summaries will not include any proprietary or confidential information.

## DISCUSSION

The course of psychotic disorders is usually characterized by relapses that severely impair an individual’s health and well-being.[3, 4] Increases in early warning signs can precede relapse by multiple weeks. However, these warning signs are highly variable between individuals and are often assessed using self-report instruments.[24] The use of language as a quantitative, objective clinical marker for the detection of early warning signs harbors great potential.[20] While prior research has demonstrated the potential of speech markers for classification of individuals as those with psychosis and those who are healthy,[39–42] for symptom severity prediction[43] and tracking,[44] as well as for predicting the conversion from a high-risk state to psychosis,[39, 45, 46] no study has prospectively evaluated their real-world applicability for relapse prediction and clinician decision support. Thus, this study aims to examine the potential of automated, AI-based transcription and speech analysis within a digital monitoring system to estimate relapse risk in individuals with psychosis.

As speech recordings are non-intrusive and non-demanding, this form of monitoring may warrant frequent sampling, allowing for weekly recordings in this study. Frequent sampling greatly improves the predictive value in longitudinal monitoring of individuals. Surveys of people with lived experiences of psychosis deemed this sampling frequency tolerable. Further, previous publications have shown that app-based symptom monitoring in individuals with psychosis is feasible.[51, 52] However, the evaluation of the feasibility and usability of a system that also includes the collection of speech data has yet to be done. For this reason, this observational study was designed to evaluate the feasibility and usability of collecting speech, self-report, and motor data across six languages using the TRUSTING app. By addressing these critical aspects, this project ensures that the digital monitoring system aligns with clinical workflows and user needs, paving the way for a future randomized controlled trial and the validation as a Clinical Decision Support System (CDSS) and Medical Device Software that aligns with the requirements of European Medical Device Regulations.

The potential benefits of this project include the early identification of relapse risk through something as natural and simple as speaking into a phone, facilitating timely interventions that may reduce distress in individuals with psychosis and might prevent hospitalization in the future. Collecting speech data in six different languages enables the assessment of the generalizability of the speech-based prediction models to diverse populations of individuals with psychosis, mitigating bias in model performance.[20] The inclusion of both individuals with psychosis and healthy participants allows for differentiation between pathological speech changes and natural linguistic variations, strengthening the reliability of automated relapse risk assessments by the monitoring system. Within the aim of explainable AI, we strive for fully interpretable and theoretically motivated models, clarifying the object and procedure of measurement, as well as the underlying rationale. By directly comparing relapse risk estimates from human raters and the digital monitoring system, the system’s trustworthiness is underscored.

As the ultimate goal is to improve the quality of life of individuals with psychosis, people with lived experience are involved in the study and monitoring system design. Through incorporating an exploratory research loop in this research, novel speech-based markers associated with relapse risk might be discovered, improving predictive accuracy, and advancing our understanding of early relapse indicators. Ultimately, the research project will generate new insights into the practicality of implementing AI-powered transcription and speech analysis in routine care, guiding the future development of scalable, user-friendly digital health solutions for relapse prevention in psychosis.

## Data Availability

Data collection has not started yet.

## Acknowledgements

We would like to thank GAMIAN-Europe and the Clinical Trials Center Zurich for the valuable collaborations.

## Author Contributions

RMH and LB drafted the initial manuscript. All authors developed the study design collaboratively. RMH, LB, GC and PH developed the statistical analysis plan. SAJ, MH, ET, KØM, LAB, EVdE, WH, and BE contributed to the development of the monitoring system. EVdE represented people with lived experience. All authors critically revised and approved the final manuscript.

## REFERENCES

1 Arciniegas DB. Psychosis. Continuum (Minneap Minn) 2015;21(3 Behavioral Neurology and Neuropsychiatry):715–36. https://journals.lww.com/continuum/fulltext/2015/06000/psychosis.15.aspx.

2 World Health Organization (WHO). Clinical descriptions and diagnostic requirements for ICD-11 mental, behavioural and neurodevelopmental disorders. Geneva: World Health Organization 2024.

3 Hui CL, Honer WG, Lee EH, et al. Predicting first-episode psychosis patients who will never relapse over 10 years. Psychol Med 2019;49(13):2206–14. doi:10.1017/S0033291718003070 [published Online First: 30 October 2018].

4 World Health Organization. World Mental Health Report: Transforming Mental Health for All, 1st edn. Geneva: World Health Organization 2022.

5 Ceraso A, Lin JJ, Schneider-Thoma J, et al. Maintenance treatment with antipsychotic drugs for schizophrenia. Cochrane Database Syst Rev 2020;8(8):CD008016. doi:10.1002/14651858.CD008016.pub3 [published Online First: 11 August 2020].

6 Neumeier MS, Homan S, Vetter S, et al. Examining Side Effect Variability of Antipsychotic Treatment in Schizophrenia Spectrum Disorders: A Meta-analysis of Variance. Schizophr Bull 2021;47(6):1601–10.

7 Correll CU, Solmi M, Veronese N, et al. Prevalence, incidence and mortality from cardiovascular disease in patients with pooled and specific severe mental illness: a large-scale meta-analysis of 3,211,768 patients and 113,383,368 controls. World Psychiatry 2017;16(2):163–80.

8 Ray WA, Chung CP, Murray KT, et al. Atypical antipsychotic drugs and the risk of sudden cardiac death. N Engl J Med 2009;360(3):225–35.

9 Murray-Thomas T, Jones ME, Patel D, et al. Risk of mortality (including sudden cardiac death) and major cardiovascular events in atypical and typical antipsychotic users: a study with the general practice research database. Cardiovasc Psychiatry Neurol 2013;2013:247486. doi:10.1155/2013/247486 [published Online First: 26 December 2013].

10 Bak M, Fransen A, Janssen J, et al. Almost all antipsychotics result in weight gain: a meta-analysis 2014.

11 Homan P, Argyelan M, Fales CL, et al. Striatal volume and functional connectivity correlate with weight gain in early-phase psychosis. Neuropsychopharmacology 2019;44(11):1948–54. doi:10.1038/s41386-019-0464-y [published Online First: 17 July 2019].

12 Kessing LV, Thomsen AF, Mogensen UB, et al. Treatment with antipsychotics and the risk of diabetes in clinical practice. Br J Psychiatry 2010;197(4):266–71.

13 Moritz S, Andreou C, Klingberg S, et al. Assessment of subjective cognitive and emotional effects of antipsychotic drugs. Effect by defect? Neuropharmacology 2013;72:179–86. doi:10.1016/j.neuropharm.2013.04.039 [published Online First: 3 May 2013].

14 La Torre A, Conca A, Duffy D, et al. Sexual dysfunction related to psychotropic drugs: a critical review part II: antipsychotics. Pharmacopsychiatry 2013;46(6):201–08. doi:10.1055/s-0033-1347177 [published Online First: 4 June 2013].

15 Moncrieff J. Antipsychotic Maintenance Treatment: Time to Rethink? PLoS Med 2015;12(8):e1001861. doi:10.1371/journal.pmed.1001861 [published Online First: 4 August 2015].

16 Kishi T, Ikuta T, Matsui Y, et al. Effect of discontinuation v. maintenance of antipsychotic medication on relapse rates in patients with remitted/stable first-episode psychosis: a meta-analysis. Psychol Med 2019;49(5):772–79. doi:10.1017/S0033291718001393 [published Online First: 18 June 2018].

17 Caseiro O, Pérez-Iglesias R, Mata I, et al. Predicting relapse after a first episode of non-affective psychosis: a three-year follow-up study. J Psychiatr Res 2012;46(8):1099–105. doi:10.1016/j.jpsychires.2012.05.001 [published Online First: 19 June 2012].

18 Davidson M, Carpenter WT. Targeted Treatment of Schizophrenia Symptoms as They Manifest, or Continuous Treatment to Reduce the Risk of Psychosis Recurrence. Schizophr Bull 2024;50(1):14–21.

19 Rubio JM, Perez-Rodriguez M. Chronic Use of Antipsychotics in Schizophrenia: Are We Asking the Right Question? Schizophr Bull Open 2022;3(1):sgac059. doi:10.1093/schizbullopen/sgac059 [published Online First: 18 October 2022].

20 Zaher F, Diallo M, Achim AM, et al. Speech markers to predict and prevent recurrent episodes of psychosis: A narrative overview and emerging opportunities. Schizophr Res 2024;266:205–15. doi:10.1016/j.schres.2024.02.036 [published Online First: 29 February 2024].

21 Herz MI, Lamberti JS, Mintz J, et al. A program for relapse prevention in schizophrenia: a controlled study. Arch Gen Psychiatry 2000;57(3):277–83.

22 Bighelli I, Rodolico A, García-Mieres H, et al. Psychosocial and psychological interventions for relapse prevention in schizophrenia: a systematic review and network meta-analysis. Lancet Psychiatry 2021;8(11):969–80. doi:10.1016/S2215-0366(21)00243-1 [published Online First: 12 October 2021].

23 Homan P, Schooler NR, Brunette MF, et al. Relapse prevention through health technology program reduces hospitalization in schizophrenia. Psychol Med 2023;53(9):4114–20. doi:10.1017/S0033291722000794 [published Online First: 30 May 2022].

24 Spaniel F, Bakstein E, Anyz J, et al. Relapse in schizophrenia: Definitively not a bolt from the blue. Neurosci Lett 2018;669:68–74. doi:10.1016/j.neulet.2016.04.044 [published Online First: 22 April 2016].

25 Eisner E, Drake R, Barrowclough C. Assessing early signs of relapse in psychosis: review and future directions. Clin Psychol Rev 2013;33(5):637–53. doi:10.1016/j.cpr.2013.04.001 [published Online First: 11 April 2013].

26 Gleeson JF, McGuckian TB, Fernandez DK, et al. Systematic review of early warning signs of relapse and behavioural antecedents of symptom worsening in people living with schizophrenia spectrum disorders. Clin Psychol Rev 2024;107:102357. doi:10.1016/j.cpr.2023.102357 [published Online First: 22 November 2023].

27 Parola A, Simonsen A, Bliksted V, et al. Voice patterns in schizophrenia: A systematic review and Bayesian meta-analysis. Schizophr Res 2020;216:24–40. doi:10.1016/j.schres.2019.11.031 [published Online First: 13 December 2019].

28 Parola A, Simonsen A, Lin JM, et al. Voice Patterns as Markers of Schizophrenia: Building a Cumulative Generalizable Approach Via a Cross-Linguistic and Meta-analysis Based Investigation. Schizophr Bull 2023;49(Suppl_2):S125-S141.

29 Palaniyappan L, Homan P, Alonso-Sánchez MF. Language Network Dysfunction and Formal Thought Disorder in Schizophrenia. Schizophr Bull 2023;49(2):486–97.

30 Surbeck W, Omlor W, Dannecker N, et al. Altered white matter microstructure of language pathways and semantic cognition deficiencies in early psychosis. Schizophrenia 2025;11(1):136.

31 Surbeck W, Hänggi J, Scholtes F, et al. Anatomical integrity within the inferior fronto-occipital fasciculus and semantic processing deficits in schizophrenia spectrum disorders. Schizophr Res 2020;218:267–75. doi:10.1016/j.schres.2019.12.025 [published Online First: 13 January 2020].

32 Cavelti M, Winkelbeiner S, Federspiel A, et al. Formal thought disorder is related to aberrations in language-related white matter tracts in patients with schizophrenia. Psychiatry Res Neuroimaging 2018;279:40–50. doi:10.1016/j.pscychresns.2018.05.011 [published Online First: 23 May 2018].

33 Cavelti M, Homan P, Vauth R. The impact of thought disorder on therapeutic alliance and personal recovery in schizophrenia and schizoaffective disorder: An exploratory study. Psychiatry Res 2016;239:92–98. doi:10.1016/j.psychres.2016.02.070 [published Online First: 2 March 2016].

34 Wang D, Gopal S, Baker S, et al. Trajectories and changes in individual items of positive and negative syndrome scale among schizophrenia patients prior to impending relapse. NPJ Schizophr 2018;4(1):10. doi:10.1038/s41537-018-0056-6 [published Online First: 20 June 2018].

35 Subotnik KL, Nuechterlein KH. Prodromal signs and symptoms of schizophrenic relapse. J Abnorm Psychol 1988;97(4):405–12.

36 Marder SR. Natural Language Processing: Its Potential Role in Clinical Care and Clinical Research. Schizophr Bull 2022;48(5):958–59.

37 Corcoran CM, Mittal VA, Bearden CE, et al. Language as a biomarker for psychosis: A natural language processing approach. Schizophr Res 2020;226:158–66. doi:10.1016/j.schres.2020.04.032 [published Online First: 1 June 2020].

38 Tang SX, Kriz R, Cho S, et al. Natural language processing methods are sensitive to sub-clinical linguistic differences in schizophrenia spectrum disorders. NPJ Schizophr 2021;7(1):25. doi:10.1038/s41537-021-00154-3 [published Online First: 14 May 2021].

39 Corcoran CM, Carrillo F, Fernández-Slezak D, et al. Prediction of psychosis across protocols and risk cohorts using automated language analysis. World Psychiatry 2018;17(1):67–75.

40 Voppel AE, de Boer JN, Brederoo SG, et al. Quantified language connectedness in schizophrenia-spectrum disorders. Psychiatry Res 2021;304:114130. doi:10.1016/j.psychres.2021.114130 [published Online First: 22 July 2021].

41 Voppel AE, de Boer JN, Brederoo SG, et al. Semantic and Acoustic Markers in Schizophrenia-Spectrum Disorders: A Combinatory Machine Learning Approach. Schizophr Bull 2023;49(Suppl_2):S163-S171.

42 de Boer JN, Voppel AE, Brederoo SG, et al. Acoustic speech markers for schizophrenia-spectrum disorders: a diagnostic and symptom-recognition tool. Psychol Med 2023;53(4):1302–12. doi:10.1017/S0033291721002804 [published Online First: 4 August 2021].

43 Rohanian M, Hüppi RM, Nooralahzadeh F, et al. Uncertainty Modeling in Multimodal Speech Analysis Across the Psychosis Spectrum 2025.

44 Palominos C, Kirdun M, Nikzad AH, et al. A single composite index of semantic behavior tracks symptoms of psychosis over time. Schizophr Res 2025;279:116–27. doi:10.1016/j.schres.2025.03.038 [published Online First: 6 April 2025].

45 Bedi G, Carrillo F, Cecchi GA, et al. Automated analysis of free speech predicts psychosis onset in high-risk youths. NPJ Schizophr 2015;1:15030. doi:10.1038/npjschz.2015.30 [published Online First: 26 August 2015].

46 Argolo F, Magnavita G, Mota NB, et al. Lowering costs for large-scale screening in psychosis: a systematic review and meta-analysis of performance and value of information for speech-based psychiatric evaluation. Braz J Psychiatry 2020;42(6):673–86.

47 Corona Hernández H, Corcoran C, Achim AM, et al. Natural Language Processing Markers for Psychosis and Other Psychiatric Disorders: Emerging Themes and Research Agenda From a Cross-Linguistic Workshop. Schizophr Bull 2023;49(Suppl_2):S86-S92.

48 Parola A, Lin JM, Simonsen A, et al. Speech disturbances in schizophrenia: Assessing cross-linguistic generalizability of NLP automated measures of coherence. Schizophr Res 2023;259:59–70. doi:10.1016/j.schres.2022.07.002 [published Online First: 1 August 2022].

49 Parola A, Jessen ET, Rybner A, et al. Vocal Markers of Schizophrenia: Assessing the Generalizability of Machine Learning Models and Their Clinical Applicability. Schizophr Bull 2025. doi:10.1093/schbul/sbaf124 [published Online First: 18 August 2025].

50 Holmlund TB, Foltz PW, Cohen AS, et al. Moving psychological assessment out of the controlled laboratory setting: Practical challenges. Psychol Assess 2019;31(3):292–303.

51 Steare T, O’Hanlon P, Eskinazi M, et al. Smartphone-delivered self-management for first-episode psychosis: the ARIES feasibility randomised controlled trial. BMJ Open 2020;10(8):e034927. doi:10.1136/bmjopen-2019-034927 [published Online First: 26 August 2020].

52 Gumley AI, Bradstreet S, Ainsworth J, et al. The EMPOWER blended digital intervention for relapse prevention in schizophrenia: a feasibility cluster randomised controlled trial in Scotland and Australia. Lancet Psychiatry 2022;9(6):477–86.

53 Bergé D, Mané A, Salgado P, et al. Predictors of Relapse and Functioning in First-Episode Psychosis: A Two-Year Follow-Up Study. Psychiatric Services 2016;67(2):227–33. doi:10.1176/appi.ps.201400316 [published Online First: 15 October 2015].

54 Bell IH, Eisner E, Allan S, et al. Methodological Characteristics and Feasibility of Ecological Momentary Assessment Studies in Psychosis: a Systematic Review and Meta-Analysis. Schizophr Bull 2024;50(2):238–65.

55 Eisner E, Bucci S, Berry N, et al. Feasibility of using a smartphone app to assess early signs, basic symptoms and psychotic symptoms over six months: A preliminary report. Schizophr Res 2019;208:105–13. doi:10.1016/j.schres.2019.04.003 [published Online First: 9 April 2019].

56 Ciampelli S, Voppel AE, Boer JN de, et al. Combining automatic speech recognition with semantic natural language processing in schizophrenia. Psychiatry Res 2023;325:115252. doi:10.1016/j.psychres.2023.115252 [published Online First: 16 May 2023].

57 Holmlund TB, Chandler C, Foltz PW, et al. Applying speech technologies to assess verbal memory in patients with serious mental illness. NPJ Digit Med 2020;3:33. doi:10.1038/s41746-020-0241-7 [published Online First: 11 March 2020].

58 Elm E von, Altman DG, Egger M, et al. The Strengthening the Reporting of Observational Studies in Epidemiology (STROBE) statement: guidelines for reporting observational studies. Lancet 2007;370(9596):1453–57.

59 Chandler C, Foltz PW, Cohen AS, et al. Machine learning for ambulatory applications of neuropsychological testing. Intelligence-Based Medicine 2020;1–2:100006.

60 Alvarez-Jimenez M, Priede A, Hetrick SE, et al. Risk factors for relapse following treatment for first episode psychosis: a systematic review and meta-analysis of longitudinal studies. Schizophr Res 2012;139(1-3):116–28. doi:10.1016/j.schres.2012.05.007 [published Online First: 1 June 2012].

61 Overall JE, Gorham DR. The Brief Psychiatric Rating Scale. Psychol Rep 1962;10(3):799–812.

62 Howes OD, Bukala BR, Chen EYH, et al. Relapse in Schizophrenia: A Systematic Review of Criteria for Clinical Studies and International Consensus Guidelines to Improve Them. Am J Psychiatry 2025;182(11):969–83. doi:10.1176/appi.ajp.20241040 [published Online First: 8 October 2025].

63 Brooke J. SUS: A “quick and dirty” usability scale. In: Jordan PW, Thomas B, Weerdmeester BA, et al., eds. Usability evaluation in industry. London: Taylor & Francis 1996:189–94.

64 Bangor A, Kortum PT, Miller JT. An Empirical Evaluation of the System Usability Scale. International Journal of Human-Computer Interaction 2008;24(6):574–94.

65 Peres SC, Pham T, Phillips R. Validation of the System Usability Scale (SUS). Proceedings of the Human Factors and Ergonomics Society Annual Meeting 2013;57(1):192–96.

66 Zhou L, Bao J, Setiawan IMA, et al. The mHealth App Usability Questionnaire (MAUQ): Development and Validation Study. JMIR Mhealth Uhealth 2019;7(4):e11500. doi:10.2196/11500 [published Online First: 11 April 2019].

67 The WHOQOL Group. The World Health Organization Quality of Life assessment (WHOQOL): Position paper from the World Health Organization. Soc Sci Med 1995;41(10):1403–09.

68 The WHOQOL Group. Development of the World Health Organization WHOQOL-BREF quality of life assessment. The WHOQOL Group. Psychol Med 1998;28(3):551–58.

69 Hall RC. Global assessment of functioning. A modified scale. Psychosomatics 1995;36(3):267–75.

70 Aas IHM. Global Assessment of Functioning (GAF): properties and frontier of current knowledge. Ann Gen Psychiatry 2010;9:20. doi:10.1186/1744-859X-9-20 [published Online First: 7 May 2010].

71 Rybarczyk B. Social and Occupational Functioning Assessment Scale (SOFAS). In: Kreutzer JS, DeLuca J, Caplan B, eds. Encyclopedia of Clinical Neuropsychology. New York, NY: Springer New York 2011:2313.

72 Rybarczyk B. Social and Occupational Functioning Assessment Scale (SOFAS). In: Kreutzer J, DeLuca J, Caplan B, eds. Encyclopedia of Clinical Neuropsychology. Cham: Springer International Publishing 2018:1.

73 Kay SR, Fiszbein A, Opler LA. The positive and negative syndrome scale (PANSS) for schizophrenia. Schizophr Bull 1987;13(2):261–76.

74 Kay SR, Opler LA, Lindenmayer JP. Reliability and validity of the positive and negative syndrome scale for schizophrenics. 01651781 1988;23(1):99–110.

75 Wistedt B, Rasmussen A, Pedersen L, et al. The development of an observer-scale for measuring social dysfunction and aggression. Pharmacopsychiatry 1990;23(6):249–52.

76 Kenward MG, Roger JH. Small Sample Inference for Fixed Effects from Restricted Maximum Likelihood. Biometrics 1997;53(3):983. http://www.jstor.org/stable/2533558.

77 Lay B, Roser P, Kawohl W. Inpatient Treatment of People With Schizophrenia: Quantifying Clinical Change Using the Health of the Nation Outcome Scales. 2632*-*7899 2021;2(1).

78 Yildiz M, Yazici A, Böke Ö. Demographic and clinical characteristics in schizophrenia: a multi center cross-sectional case record study. Turk Psikiyatri Derg 2010;21(3):213–24.

79 Eurostat. In-patient average length of stay (days): Eurostat 2022.

80 Haynes A, Lenz A, Stalder O, et al. presize: An R-package for precision-based sample size calculation in clinical research. JOSS 2021;6(60):3118.

81 Hanley JA, McNeil BJ. The meaning and use of the area under a receiver operating characteristic (ROC) curve. Radiology 1982;143(1):29–36.

82 Harrell FE. Regression modeling strategies: With applications to linear models, logistic and ordinal regression, and survival analysis. Cham, Heidelberg, New York: Springer 2015.

83 World Medical Association. World Medical Association Declaration of Helsinki: ethical principles for medical research involving human subjects. JAMA 2013;310(20):2191–94.

84 Global Alliance of Mental Illness Advocacy Networks-Europe (GAMIAN-Europe). TRUSTING 2025. Available at: https://www.gamian.eu/trusting/ Accessed October 30, 2025.

